# Group IIA Secreted Phospholipase A_2_ Plays a Central Role in the Pathobiology of COVID-19

**DOI:** 10.1101/2021.02.22.21252237

**Authors:** Justin M. Snider, Jeehyun Karen You, Xia Wang, Ashley J Snider, Brian Hallmark, Michael C. Seeds, Susan Sergeant, Laurel Johnstone, Qiuming Wang, Ryan Sprissler, Hao Helen Zhang, Chiara Luberto, Richard R. Kew, Yusuf A Hannun, Charles E. McCall, Guang Yao, Maurizio Del Poeta, Floyd H. Chilton

## Abstract

There is an urgent need to identify cellular and molecular mechanisms responsible for severe COVID-19 disease accompanied by multiple organ failure and high mortality rates. Here, we performed untargeted/targeted lipidomics and focused biochemistry on 127 patient plasma samples, and showed high levels of circulating, enzymatically active secreted phospholipase A_2_ Group IIA (sPLA_2_-IIA) in severe and fatal COVID-19 disease compared with uninfected patients or mild illness. Machine learning demonstrated that sPLA_2_-IIA effectively stratifies severe from fatal COVID-19 disease. We further introduce a PLA-BUN index that combines sPLA_2_-IIA and blood urea nitrogen (BUN) threshold levels as a critical risk factor for mitochondrial dysfunction, sustained inflammatory injury and lethal COVID-19. With the availability of clinically tested inhibitors of sPLA_2_-IIA, our study opens the door to a precision intervention using indices discovered here to reduce COVID-19 mortality.

## Introduction

Host resistance and disease tolerance are paramount to mounting a successful defense against infections such as SARS-CoV-2^1^. Up to 80% of individuals infected with SARS-CoV-2 are asymptomatic or develop mild to moderate symptoms. However, others progress to severe and life-threatening conditions, requiring hospitalization and specialized medical care. COVID-19 severity correlates with respiratory symptoms (i.e. dyspnea, hyperpnea, hypoxemia, pulmonary infiltration) and concomitant multiple organ failure with disseminated intravascular coagulation^2^. Consequently, there is an urgent need to elucidate central molecular mechanisms that underly severe and fatal COVID-19 disease to develop targeted therapeutic approaches.

Early studies suggested that the host response to COVID-19 may be associated with an excessive proinflammatory response caused by a cytokine storm syndrome (CSS)^3-5^. However, more recent studies show that a persistent CSS is uncommon (3-4%) in severe COVID-19 disease, where high-dose steroids benefit only a small proportion of individuals with organ failure^6-8^. Mounting evidence supports that immunometabolic suppression and not CSS compromises host immunity, leading to unrestrained viral replication and severe COVID-19^9,10^. Even when viral burdens are reduced, immunopathologies including tissue and organ damage often remain^11-13^.

Lipid metabolism plays an important role in determining COVID-19 outcomes. Early lipidomic studies^14,15^ revealed that severe COVID-19 modifies the circulating lipidome, with decreases in plasma levels of phospholipids and elevated quantities of lyso-phospholipids (lyso-PL), unesterified unsaturated fatty acids (UFA), and acylcarnitines. This lipidomic pattern suggests that increased COVID-19 severity may be accompanied by cellular or circulating phospholipase(s) that cleave intact phospholipids from cellular and mitochondrial membranes to form lyso-PL and UFA. Among phospholipases, the secreted phospholipase A_2_ (sPLA_2_) family includes 12 members with highly conserved characteristics, including low molecular weight (13-17 kDa), high Ca^2+^ levels for catalytic activity, and the presence of histidine/aspartic acid dyads in the catalytic site^16^. sPLA_2_-IIA elevations occur under various clinical conditions, including sepsis and systemic bacterial infections, adult respiratory disease syndrome (ARDS), atherosclerosis, cancer, and multiple organ trauma.^16^ Basal levels of circulating sPLA_2_-IIA in healthy humans are ∼3 ng/ml; however, sPLA_2_-IIA plasma concentrations can reach 250-500 ng/ml during acute sepsis^17^.

Here, we identify lipidomic signatures of PLA_2_ hydrolysis and mitochondrial dysfunction that correspond with COVID-19 severity in 127 patient plasma samples. Extremely high levels of circulating sPLA_2_-IIA mirrored disease severity, particularly in deceased COVID-19 patients. Circulating sPLA_2_-IIA remained catalytically active and was associated with indices of disease severity, hyperglycemia, kidney dysfunction, hypoxia, anemia, and multiple organ dysfunction. Importantly, unbiased machine learning identified sPLA_2_-IIA as a central node in predicting survivors from non-survivors in severe COVID-19 cases. Our findings demonstrate that the plasma sPLA_2_-IIA level, together with BUN, may serve as a potent clinical indicator for COVID-19 related mortality and suggest that the use of sPLA_2_-IIA inhibitors may provide a novel, targeted therapeutic approach to treat severe COVID-19 disease.

## Results

### Patients

A total of 127 patient plasma samples collected between May and July 2020 were analyzed. The demographics and baseline clinical characteristics of the patients are shown in Table 1. Age differed across groups, with deceased COVID-19 patients being older on average (Figure S1). There were no significant trends in BMI or obesity. Prevalence of various co-morbidities was comparable across groups, except for rheumatologic disease in mild COVID-19 patients (Figure S1). Severe and deceased COVID-19 patients experienced more complications, with higher incidences of cardiac arrest, acute kidney injury/renal failure, bacterial pneumonia, ARDS, and sepsis (Figure S1).

**Table 1:**
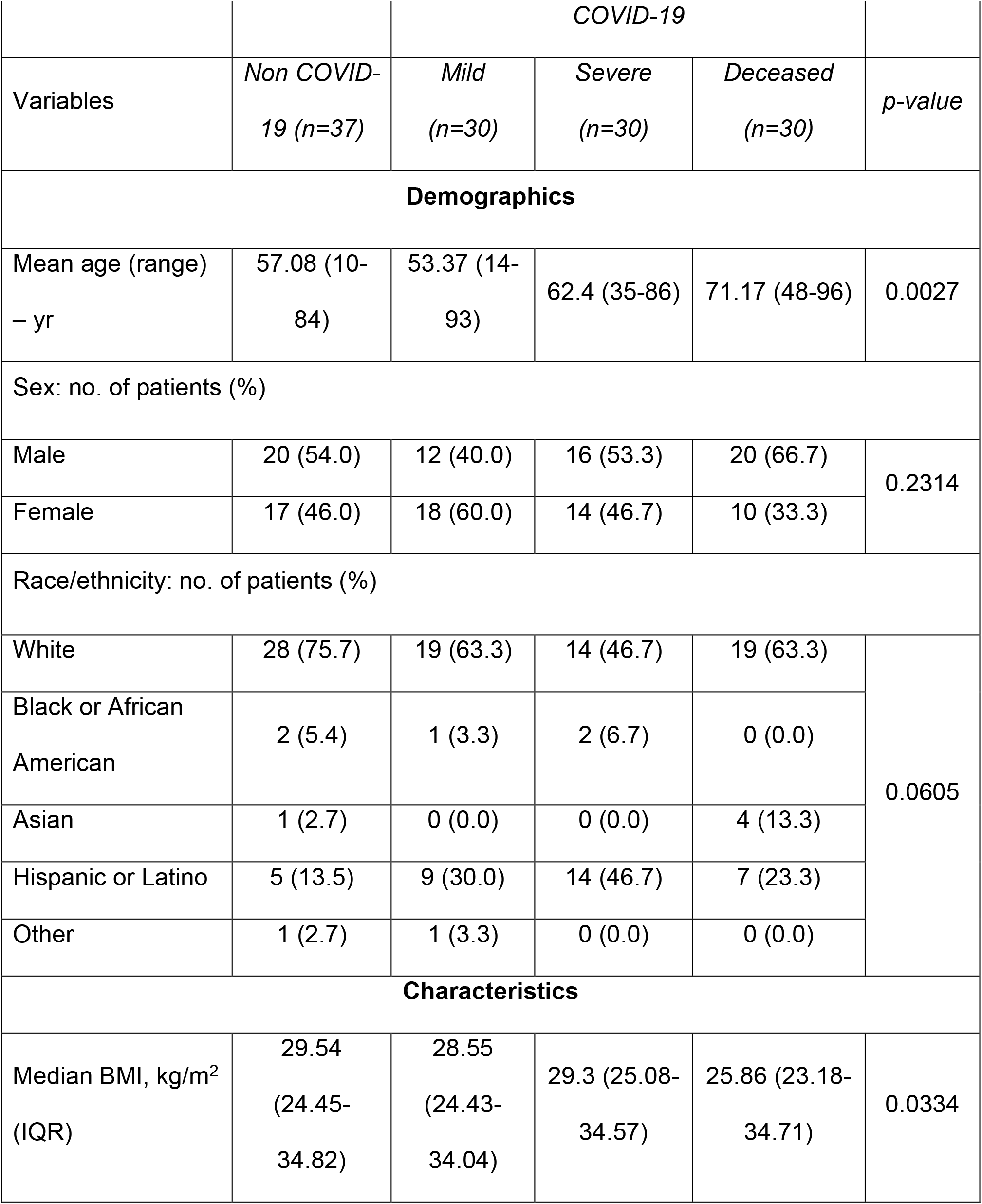
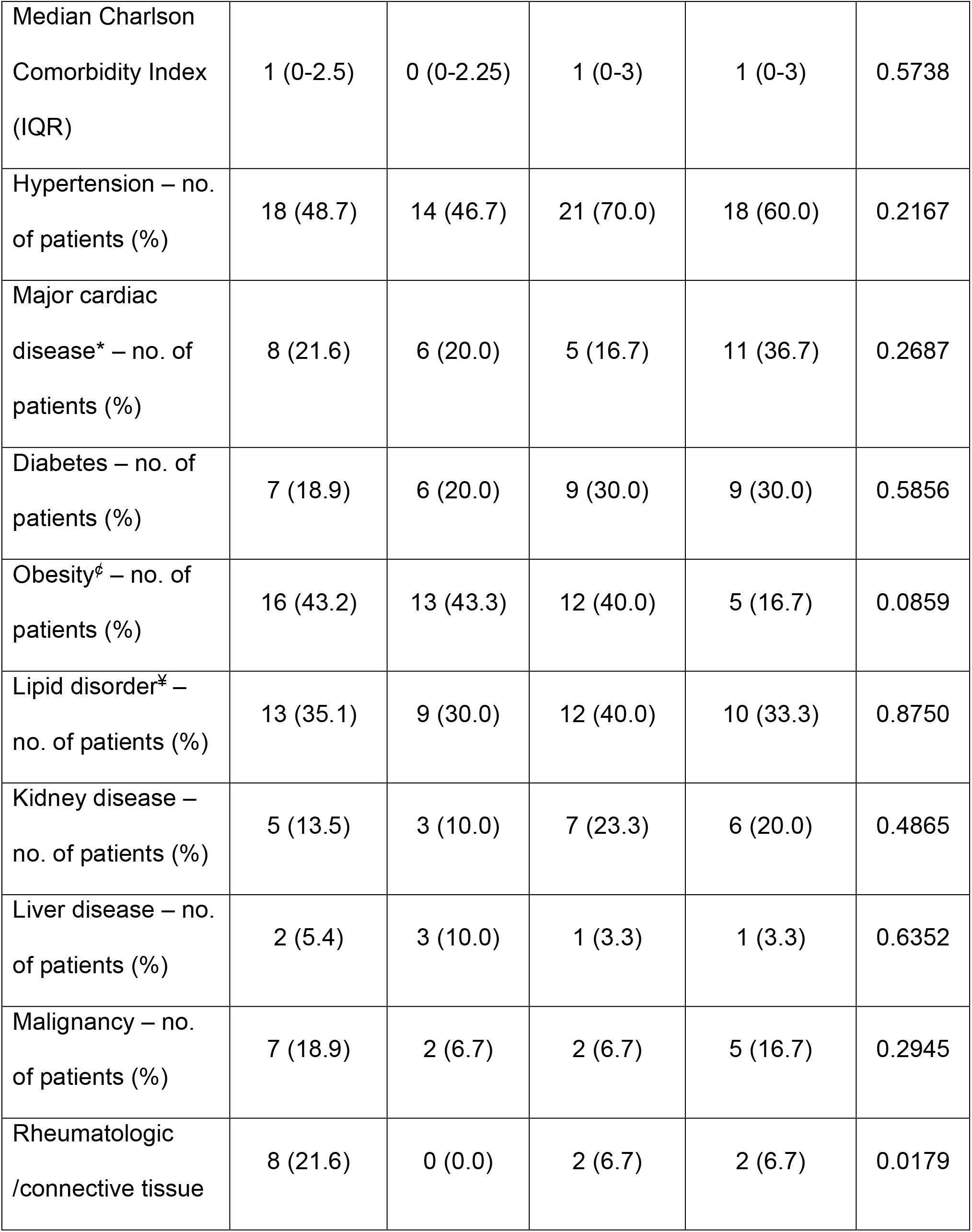
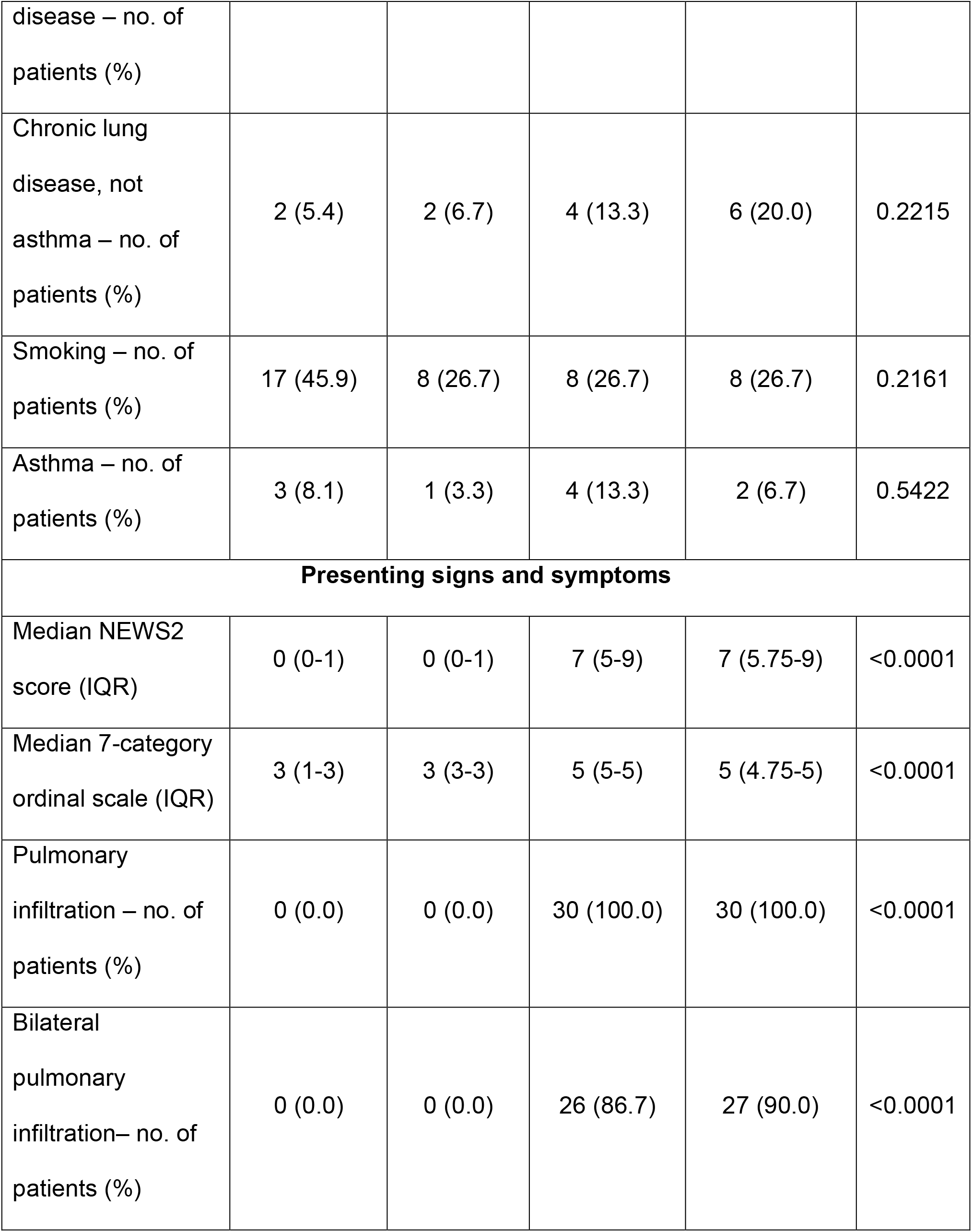
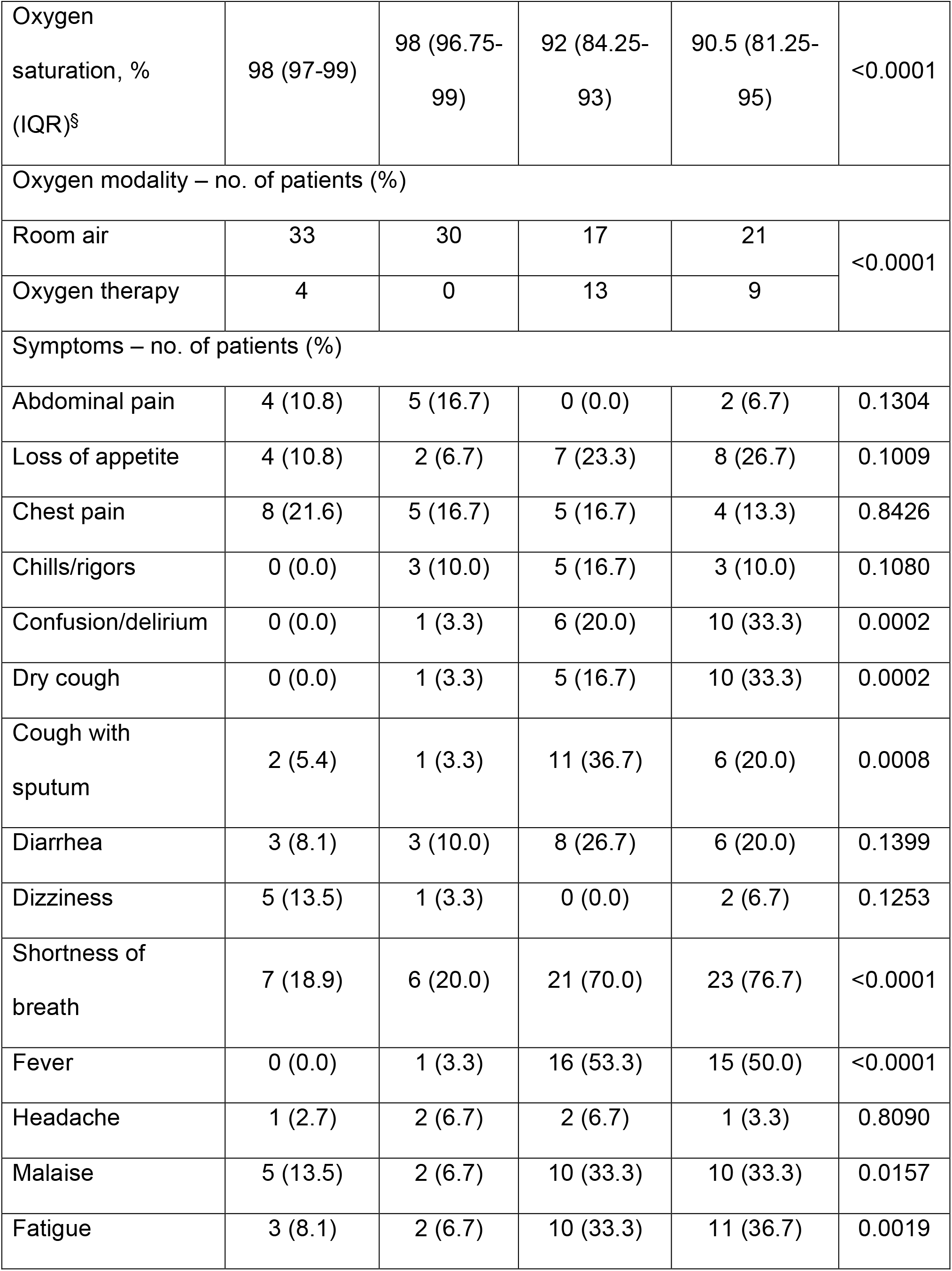
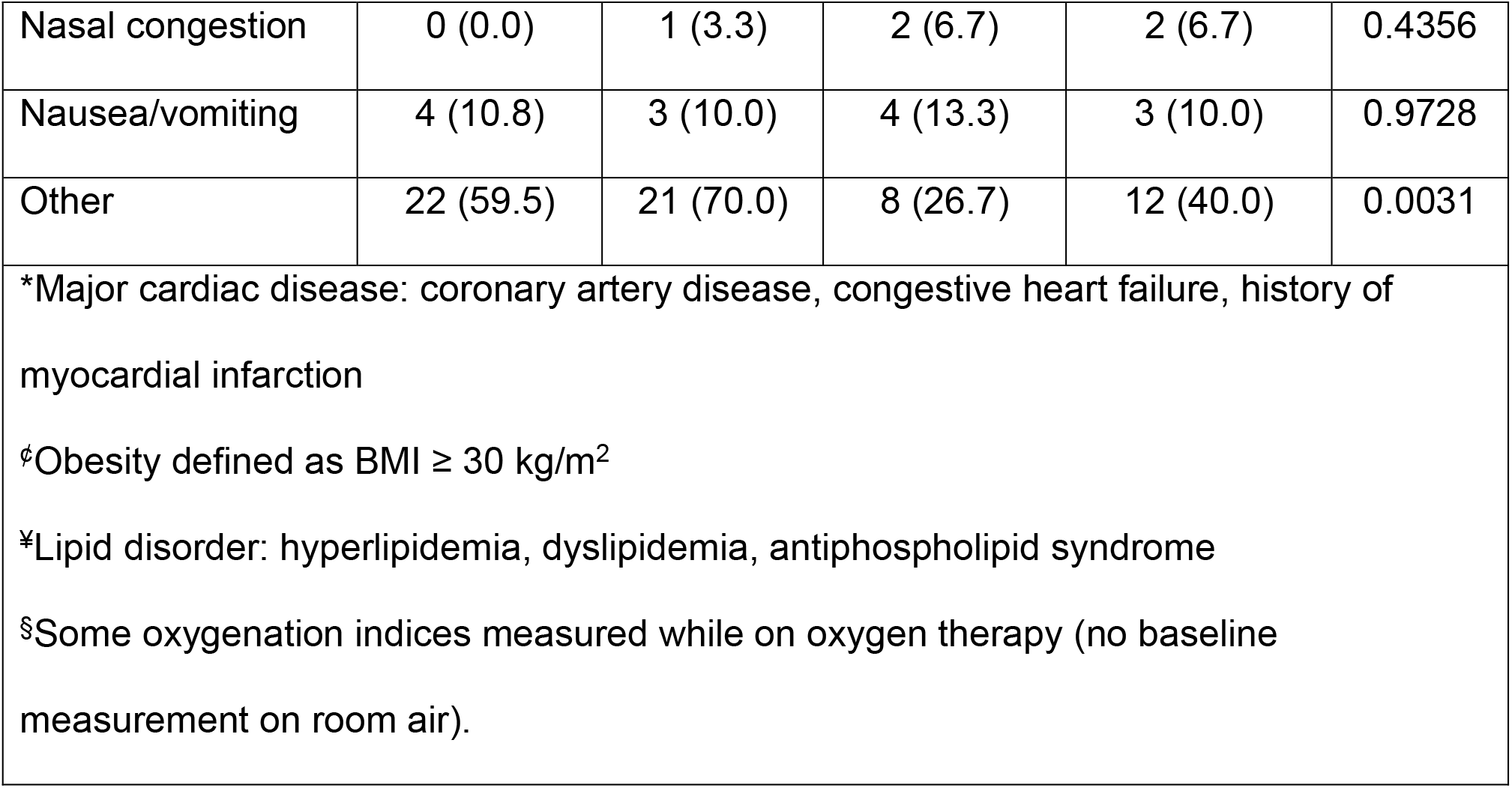
Demographics and Clinical Characteristics at Baseline. All categorical variables are represented as proportions (%) whereas continuous variables are reported as median (interquartile range). D’Agostino-Pearson normality test was used to assess continuous variables and determined all that had non-Gaussian distributions; Kruskal-Wallis test were then used to assess for equality of group variance. Categorical variables were compared using the chi-square test. P-values reflect comparisons of group variance; significant trends are reported in Figure S1.

### Plasma Lipidomic Profiles and Covid Disease Status

Untargeted lipidomic analysis of the plasma samples revealed that the most significant changes in the lipid profile occurred in deceased COVID-19 patients (Figure 1A), with 181 unique molecules identified. Further analysis of the 20 most significant molecules demonstrated an enrichment in metabolites associated with acylcarnitine and phospholipid metabolism (Figure 1B). Initial analysis showed that several lyso-phosphatidylethanolamine (lyso-PE) molecular species typified by C16eLysoPE and unsaturated fatty acids such as linoleic (18:2) and oleic acids (18:1) were elevated in severe/deceased COVID-19 patients (Figure 1C). Targeted lipidomics confirmed the compositional untargeted lipidomic analysis, showing significant increases in major molecular species of lyso-PE and lyso-phosphatidylserine (lyso-PS) while demonstrating no changes in lyso-phosphatidylcholine (lyso-PC) (Figure S2). Together, this suggested hydrolysis by a PLA_2_ activity (Figure 1D).

**Figure 1.**
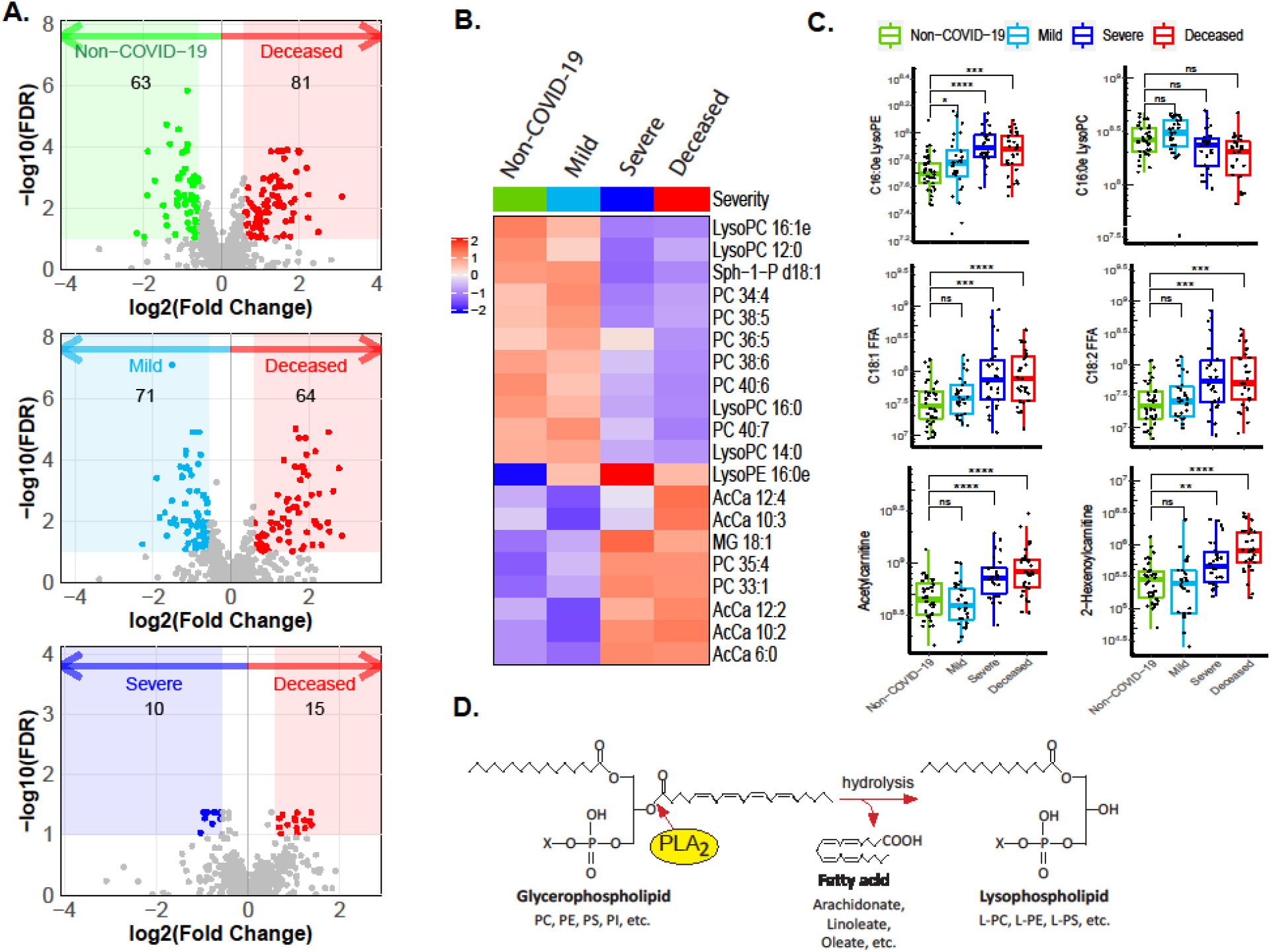
Untargeted Lipidomic Analysis and COVID-19 Status. Plasma samples from non-COVID-19, mild, severe and deceased COVID-19 patients were subjected to untargeted metabolomics analyses. Lipidome data were extracted from the metabolomics data set and analyzed. A) Volcano plots show significant alterations in the lipidome of the deceased COVID-19 patients compared with non-COVID19, mild and severe COVID-19 patients. B) Heatmap of the top 20 metabolites whose abundances varied significantly across non-COVID-19, mild, severe, and deceased COVID-19 patients. C) Abundances of two lyso-PL, two free fatty acids (FFA) and two short chain acyl carnitines extracted from the untargeted lipid data were calculated and analyzed using a one-sided Wilcoxon test. Significance indicated as: * p<0.05; ** p<0.01; *** p<0.001; **** p<0.0001. D) Model of PLA_2_ reaction showing how PLA_2_ hydrolyzes the sn-2 position of the glycerol backbone of phospholipids to form lyso-PL and FFA products.

Plasma short chain acyl carnitine and particularly acetylcarnitine has recently been shown to serve as an independent prognostic biomarker for mortality in sepsis and heart failure^18,19^. Interestingly, short- and medium-chain acylcarnitines (acetyl and hexanoyl carnitines) were also elevated in severe and deceased COVID-19 patients (Figure 1C). Furthermore, acetylcarnitine showed high areas under ROC curves: 0.810 (95% CI, 0.694-0.925) for mild vs. severe and 0.849 (95% CI, 0.752-0.945) for mild vs. deceased (Figure S3A). Additionally, plasma concentrations of mitochondrially encoded cytochrome B (MT-CYB) and cytochrome c oxidase subunit III (MT- COX3) were significantly elevated in deceased COVID-19 patients compared to non-COVID-19 and mild COVID-19 patients, suggesting elevated mtDNA (Figure S3B).

### Circulating Secreted PLA_2_-IIA Associated with Covid-19 Disease Status

Given the critical role of sPLA_2_-IIA in several related diseases^16^, its levels were quantified in all 127 patients. Figure 2A shows the distribution of sPLA_2_-IIA in all patients and the marked increase of sPLA_2_-IIA in severe (66.6 ± 25.2 ng/ml) and deceased COVID-19 patients (187.3 ± 46.6 ng/ml) compared to non-COVID-19 (24.1 ± 7.4 ng/ml) and mild COVID-19 patients (31.5 ± 9.4 ng/ml). There was heterogeneity among the severe COVID-19 patients with 48% of severe patients having relatively normal (< 10 ng/ml) levels of circulating sPLA_2_-IIA levels (inset, Figure 2A). In contrast, sPLA_2_-IIA in all deceased COVID-19 patients exceeded 10 ng/ml, with 46% of these patients having at least 10-fold higher levels (ranging from 102 ng/ml to 1020 ng/ml). Enzymatic assays showed sPLA_2_-IIA was catalytically active (Figure 2B), with a strong correlation (r^2^ = 0.84, p = 1.2 x 10^−13^) between sPLA_2_-IIA levels and enzymatic activity (Figure 2C).

**Figure 2.**
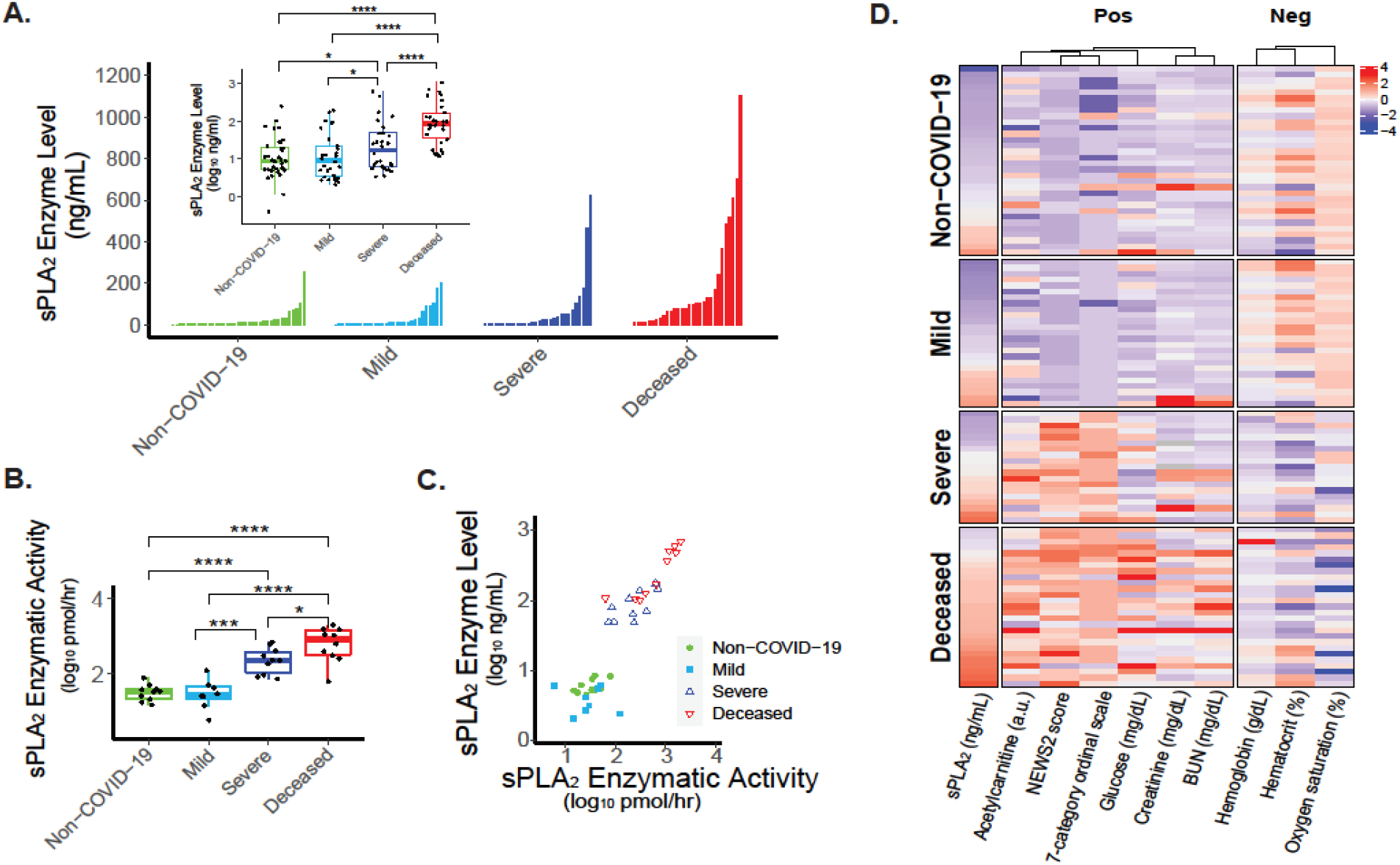
Association of sPLA_2_-IIA and COVID-19 Status. A) sPLA_2_-IIA levels were determined in 127 plasma samples. The inset boxplot demonstrates the medians, bottom and top quantiles, and statistical significance across all four groups. B) sPLA_2_ enzymatic activity was assayed within the plasma of selected samples (see Supplementary Methods). C) The scatter plot depicts a linear relationship between plasma sPLA_2_-IIA levels and sPLA_2_ activity. D) The heatmap shows the significant associations between sPLA_2_-IIA levels and clinical indices of disease severity (FDR < 0.05 in Spearman correlation, samples with > 15 missing values were filtered out). Color scheme: intensity representing the magnitude of value (mean-centered, scaled by the standard deviation, and log-transformed with non-Gaussian distribution).

Elevated levels of plasma sPLA_2_-IIA were significantly associated with several critical clinical indices (Figure 2D). Its positive correlation with higher baseline NEWS2 and 7-category ordinal scale scores suggests a role of sPLA_2_-IIA in disease severity. The positive correlation of sPLA_2_- IIA with glucose levels highlights its link to inflammation. Consistently, hyperglycemia has been reported to be an important prognostic factor for COVID-19, associated with a pro-oxidative/proinflammatory state^20^. The positive correlations with creatinine and BUN levels demonstrate how sPLA_2_-IIA levels may also reflect kidney dysfunction. Finally, the negative correlations with hematocrit, hemoglobin levels, and baseline oxygen saturation suggest elevated sPLA_2_-IIA levels may be associated with hypoxia, anemia, and multiple organ dysfunction^21^.

### Levels of Secreted PLA_2_-IIA as a Central Predictor of Covid-19 Mortality

The eighty clinical indices measured in our cohort of 127 patients were analyzed by machine learning models. First, a decision tree was generated by recursive partitioning to identify critical indices that separate the four patient groups with high accuracy (area under ROC curve = 0.93- 1.0, Figure 3A inset). Specifically, patients positive for COVID-19 were stratified using the predictor “7-category ordinal scale” into “mild” and “severe or deceased”, with 91% and 100% accuracy, respectively. Of patients in “severe or deceased”, sPLA_2-_IIA levels were found to effectively separate the survivors from non-survivors. Those with sPLA_2-_IIA <10 ng/mL were classified as “severe” but not “deceased” with a 100% accuracy; in contrast, those with sPLA_2-_IIA ≥ 10 ng/mL were placed as “deceased” with a 63% accuracy. Of these sPLA2-high (≥ 10 ng/mL) patients, BUN levels further helped improve the prediction of survival: those with BUN <16 mg/dL were classified as “severe” but not “deceased” (100% accuracy); conversely, those with BUN ≥ 16 mg/dL were markedly enriched (76%) with “deceased” patients. In short, the decision tree identified sPLA_2_ and BUN as two critical risk factors for COVID-19 mortality. Correspondingly, the effective separation of mild, severe, and deceased COVID-19 patients can be visualized in the sPLA2-BUN boundary graphs (Figure 3B).

**Figure 3.**
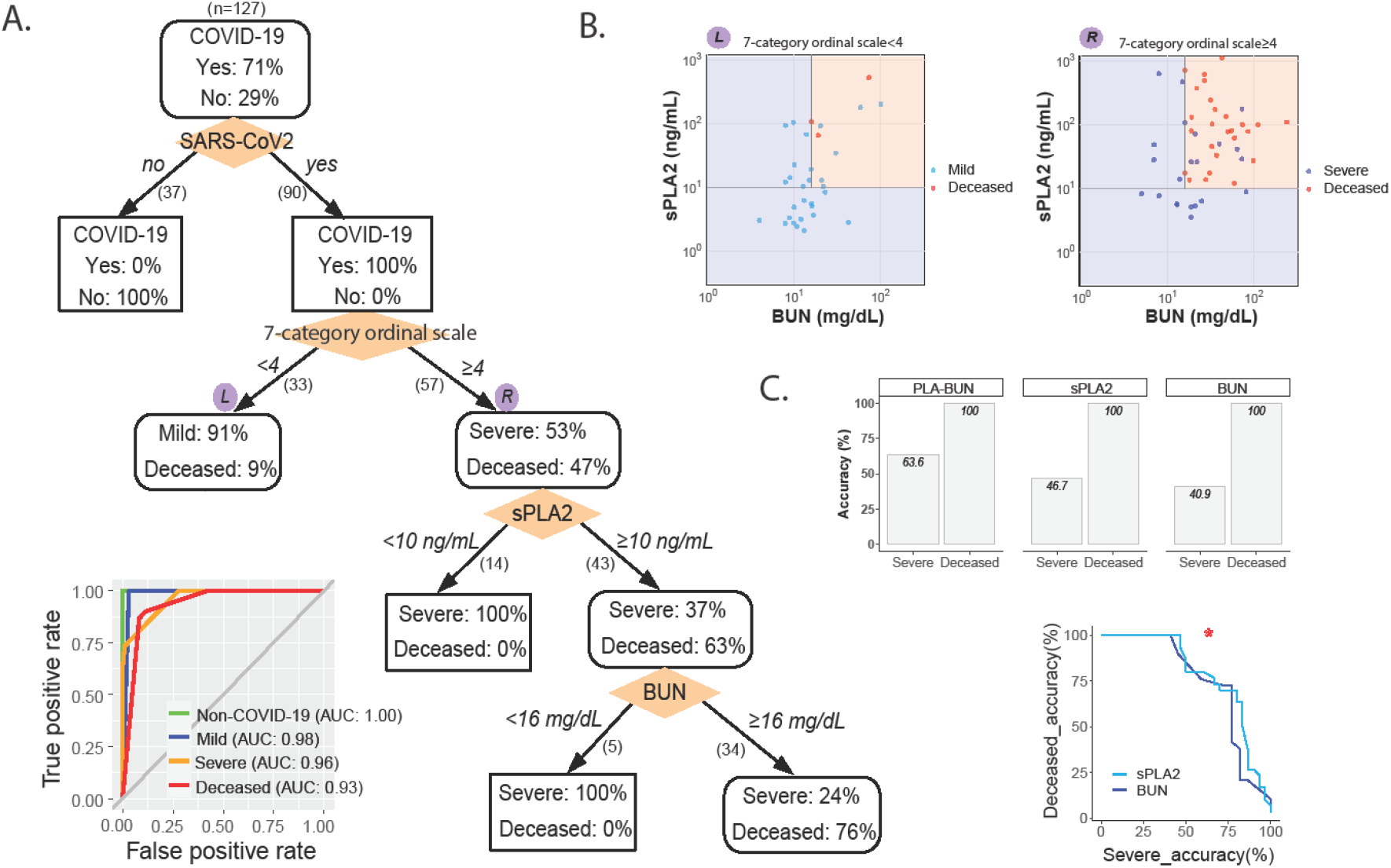
A Clinical Decision Tree Predicting COVID-19 Severity and Mortality. A) *The tree model*. Patients are classified based on the indicated clinical indices (shown in orange diamonds) and boundary conditions (above split arrows). The number of patients following each split is shown in parentheses beneath the split arrow (patients with missing index values were not split). In each node, percentages of patients in corresponding categories are shown. (Inset) The area under the ROC curve, AUC, of the tree in determining each group membership (e.g., deceased vs. non-deceased). B) Decision surface based on the sPLA_2_ and BUN boundary conditions in A. Left (L) and right (R) graphs show the results of applying the sPLA2 and BUN boundary conditions to the L and R subsets of patients (split following the 7-category ordinal scale), as indicated in A. C) The PLA-BUN index. (Top) The accuracies of combining both decision boundary conditions of sPLA2 and BUN as in B (i.e., the PLA-BUN index) in classifying severe and deceased COVID-19 patients were compared to such classification accuracies using the single decision boundary (as in B) of either sPLA2 or BUN. (Bottom) The accuracies of the PLA-BUN index in classifying severe (x-axis) and deceased (y-axis) COVID-19 patients are indicated with a red star, which are higher than such classification accuracies of using the single index of sPLA2 (light blue curve) or BUN (dark blue curve) with varying cutoff values in the corresponding data range (sPLA2, 3.4-1101.2 ng/mL; BUN, 5-242 mg/dL).

The decision tree provides a clinical blueprint to identify COVID-19 patients that progress to mortality. To validate sPLA_2_ and BUN as two critical predictors of COVID-19 mortality, an additional random forest analysis was performed to evaluate the relative importance of all 80 clinical indices. We randomly selected subsets of patients and features (clinical indices) and built decision trees (1,000 trees each in 10 repeats) to provide a robust assessment of feature importance in separating severe vs. deceased COVID-19 patients. Consistently, sPLA_2_ and BUN were identified as the top 2 features ranking significantly higher (p < 0.0001) above all other clinical indices to accurately predict COVID-19 related mortality (Figure 4). Importantly, combining both decision boundary conditions of sPLA_2_ and BUN (the PLA-BUN index) performed more accurately than using either index alone (Figure 3C).

**Figure 4.**
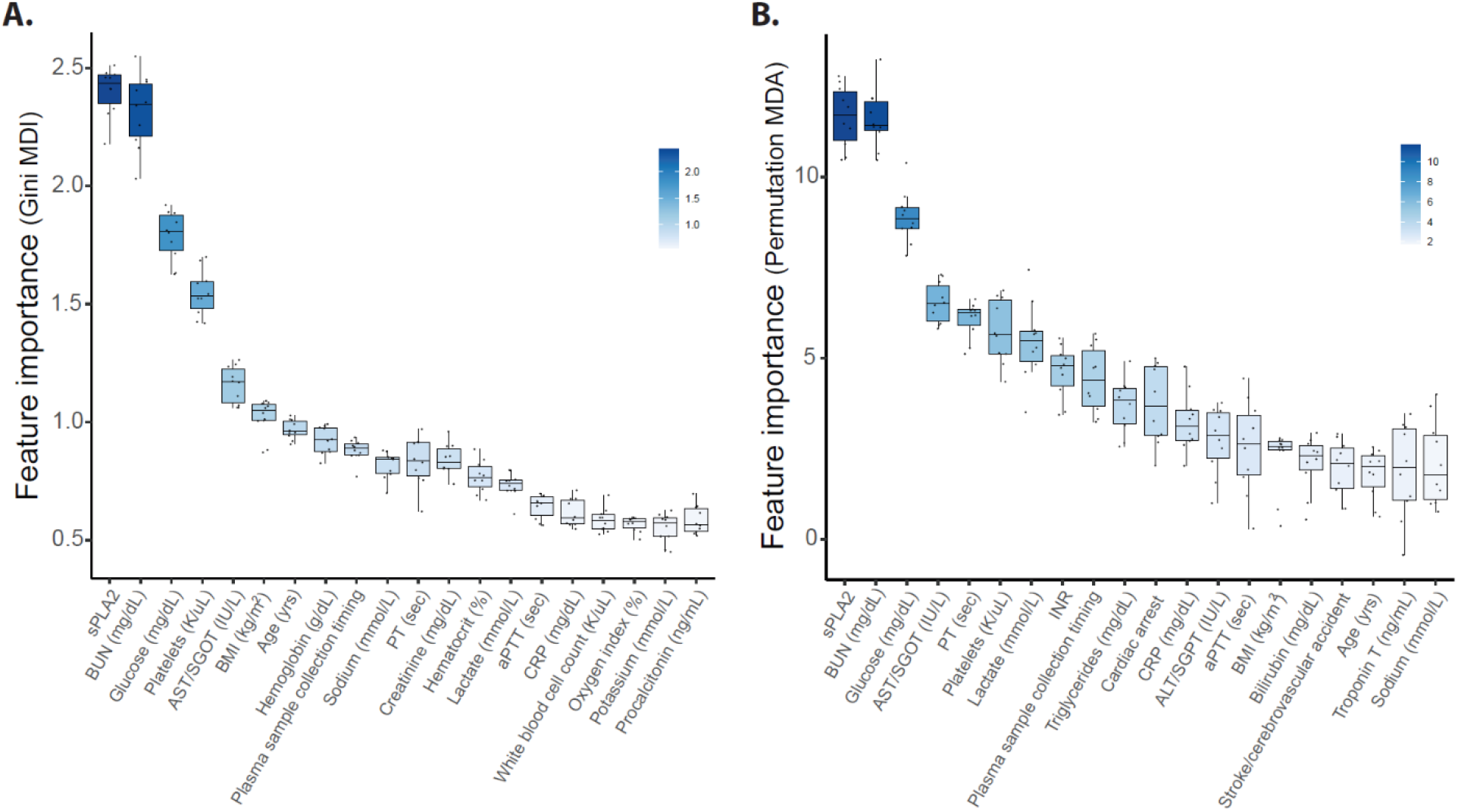
Feature importance ranking of clinical indices. The relative importance of the 80 clinical indices in separating the deceased from severe COVID-19 patients (n = 30 each) was evaluated in a random forest analysis (tree number = 1,000 each in 10 repeats). The importance of a feature (i.e., clinical index) was assessed by the decrease of prediction accuracy when such a feature was excluded from the model, based on the Gini impurity following a node split (A; MDI, Mean Decrease Impurity) and the permuted values of the feature (B; MDA, Mean Decrease Accuracy). The top 30 features in each importance measurement are shown (color scheme is proportional to the importance score).

## Conclusion

Given growing evidence suggesting that lipid metabolism plays a critical role in determining COVID-19 outcomes, we sought to identify molecular mechanisms that reconcile key lipidomic changes. Untargeted lipidomic analysis in this study, consistent with other reports, suggest that PLA_2_ activation and mitochondrial dysfunction are central determinants of COVID-19 severity and mortality^14,15,22,23^. Specifically, significant elevations in lyso-PLs (PE, PS, but not PC) and increased linoleic and oleic acid levels are hallmarks of catalysis by a sPLA_2_ isoform. Further PLA_2_ analysis revealed that high concentrations of catalytically active sPLA_2_ circulate in the plasma of severe and deceased COVID-19 patients. Elevated PLA_2_ activity in plasma of severe sepsis patients was initially described in the 1980s^24-27^. These studies showed that high levels of plasma PLA_2_ activity (∼20-fold increase) were not only sustained, but also further elevated in patients who succumbed to sepsis, suggesting distinct cellular and molecular mechanisms associated with mortality. In the present study, PLA_2_ activity was identified as sPLA_2_-IIA^28^, and deceased COVID-19 patients averaged 18.7-fold higher than normal (<10 ng/ml) with concentrations as high as 1,020 ng/ml. Forty-six percent of deceased COVID-19 patients had concentrations 10-fold or greater, and elevated sPLA_2_-IIA levels in patients were associated with several indices of COVID-19 disease severity (e.g., hyperglycemia, kidney dysfunction, hypoxia, anemia, and multiple organ dysfunction). These findings support that sPLA_2_-IIA is a central mediator in determining poor COVID-19 outcomes.

Further evidence of lipid dysregulaton during systemic inflammatory disease were the elevations in circulating short-chain acylcarnitines (particularly acetylcarnitine) and mtDNA in severe and deceased COVID-19 patients indicating systemic mitochondrial energy derangement and defective fatty acid oxidation^29,30^. Elevated plasma acetylcarnitine is not only associated with multiple organ failure, inflammation, and infection in sepsis patients, but is an important predictor of mortality^18^. Our data not only corroborates emerging evidence of elevated acetylcarnitine in COVID-19 patients, but also show that acetylcarnitine is an indicator of COVID-19 related mortality. Increases in mtDNA levels further support severe disease and mortality in COVID-19^31^. Together, our data indicate that defective fatty acid oxidation and mitochondrial dysfunction within vital organs may not only induce inflammatory damage^32,33^, but could underly sPLA_2_-IIA-related COVID-19 severity and mortality.

The clinical decision tree developed in this study offers a framework to identify COVID-19 patients at high risk for progressing to mortality (Figure 3A). Circulating sPLA_2_-IIA ≥ 10 ng/ml was identified as a critical marker to stratify deceased from severe (yet survived) COVID-19 patients, pinpointing sPLA_2_-IIA as a risk factor for COVID-19 related mortality. Our decision tree further corroborates BUN as an important risk factor associated with COVID-19 mortality^34^. Strikingly, sPLA_2_-IIA and BUN also stood out as the two unique and essential predictors of the mortality in severe COVID- 19 patients, with their feature importance rankings significantly higher (p < 0.0001) than other clinical indices in our random forest analysis. When we combined sPLA_2_-IIA and BUN into a PLA- BUN index, severe and deceased patients separated more accurately than using either one alone. Thus, we introduce the PLA-BUN index as a novel and potentially powerful clinical tool to predict COVID-19 related mortality and stratify severe patients to receive treatment of sPLA_2_-IIA inhibitors.

sPLA_2_-IIA has direct and organism-wide pathogenic characteristics (Figure 5)^16,35-38^ with the capacity to impact COVID-19 severity and patient outcomes. During cell activation and initiation of multiple cell death mechanisms, anionic phospholipids PS and PE are externalized, exposing them to phospholipid hydrolysis by sPLA_2_-IIA (Figure 5A)^39^. Hydrolysis of cellular membranes would broadly invoke tissue damage and organ cell dysfunction. Additionally, activated cells and damaged tissues/organs secrete extracellular mitochondria^35^. As mitochondrial phospholipids are the preferred substrates for sPLA_2_-IIA, our data supports that catalysis releases mtDNA, acetylcarnitine and several danger-associated molecular patterns (DAMPs) (Figure 5B)^40^. Damaged mitochondria can then be internalized by bystander leukocytes (Figure 5C) to increase inflammatory mediators including lyso-PLs, unsaturated fatty acids, eicosanoids, and cytokines. sPLA_2_-IIA also hydrolyzes platelet-derived extracellular vesicles (EV) to release cyclooxygenase, thromboxane synthase, and 12-lipoygenase inflammatory eicosanoids^16^. Collectively, this study suggests that cell injury and destructive events spreading across organs amplify inflammation with the potential to further damage tissues and organs in severe and fatal COVID-19 disease.

**Figure 5.**
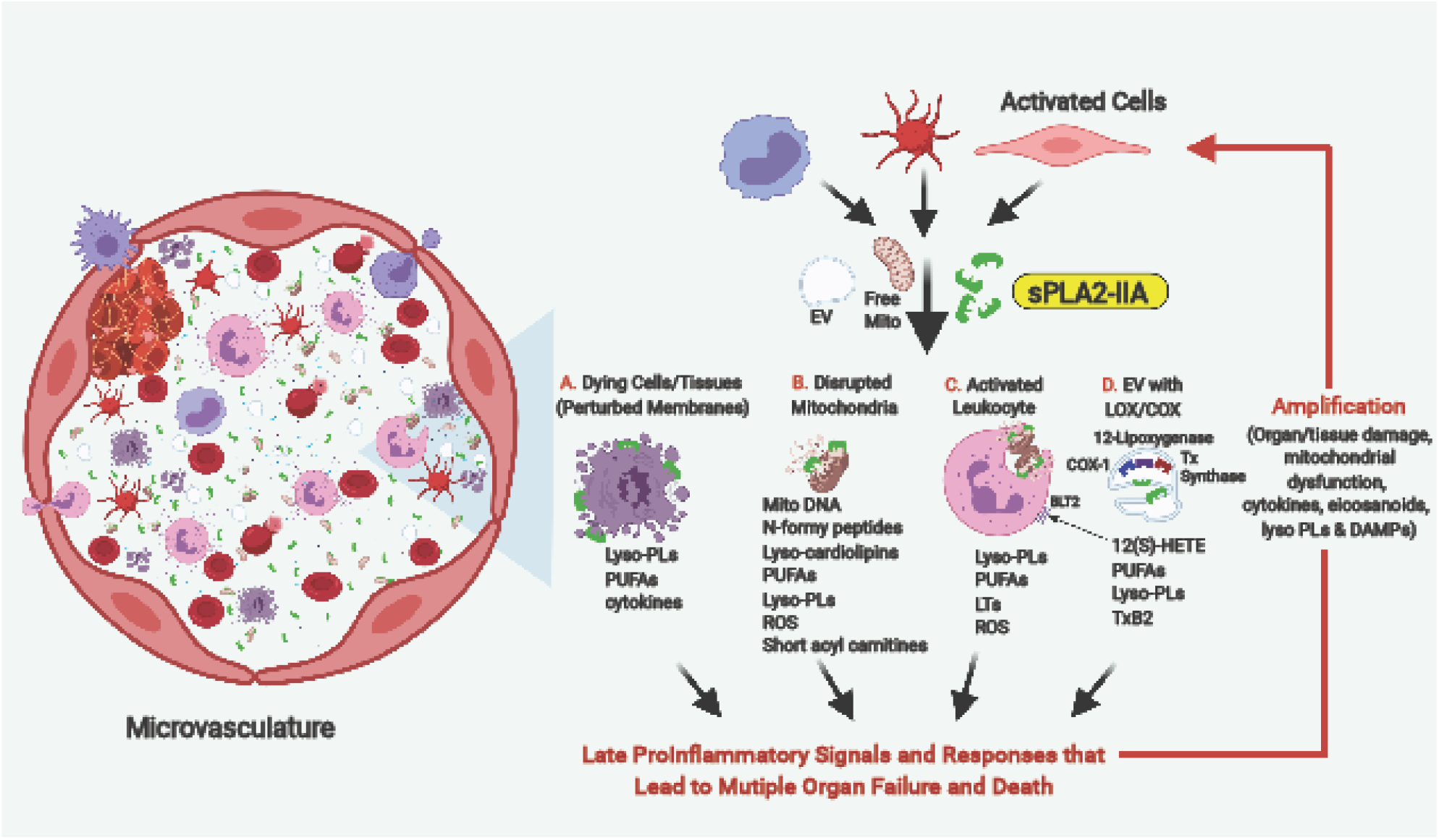
Potential Central Roles of sPLA2-Group II in Mediating COVID-19 Severity and Outcomes. A) sPLA_2_-IIA hydrolyzes phospholipids in activated and dying cells within tissues and organs. B) sPLA_2_-IIA hydrolyzes mitochondrial membranes. C) Damaged mitochondria are internalized by bystander leukocytes. D) sPLA_2_-IIA hydrolyzes extracellular vesicles (EV) containing eicosanoid-producing enzymes (cyclooxygenase (COX-1), thromboxane synthase (Tx synthase) and 12-lipoxygenase). Collectively, these events amplify late inflammatory responses, further damaging tissues and organs. Progressive tissue/organ damage plays a critical role in progressing COVID-19 disease to mortality.

As a retrospective observational study, there are limitations. Since study patients were selected on plasma sample availability, the study is subject to potential confounders and may not represent the general population. Additionally, clinical data availability was restricted to existing medical records, and there were missing values in the dataset. Furthermore, temporal relationships are difficult to assess as plasma sampling was not standardized. Finally, given the chaotic nature of COVID-19 management in early 2020, current standards of care may differ. In spite of these limitations, the study provides key mechanistic insights into COVID-19 mortality. It identifies sPLA_2_-IIA as a previously unrecognized and plausible life-threatening mechanism underlying COVID-19 severity and mortality. It also provides a clinical blueprint to identify those COVID-19 patients at risk of death and supports sPLA_2_-IIA as a therapeutic target.

sPLA_2_-IIA secretion increases during other forms of critical illness, often complicated by multiple organ failure and high mortality rates^41-43^. Consequently, structurally diverse classes of sPLA_2_-IIA inhibitors were developed that selectively inhibit sPLA_2_-IIA. Although deemed safe for clinical use, clinical trials using a sPLA_2_-IIA inhibitor only improved survival in sepsis patients when treatment was initiated within 18 hours of organ failure^44,45^. Further examination of these studies’ design revealed limitations: 1) patient selection criteria did not incorporate patient sPLA_2_-IIA levels and 2) circulating sPLA_2_-IIA levels were not reported in the studies. Therefore, inappropriate patient selection likely contributed to patient heterogeneity, resulting in negative findings. Indeed, a recent study reported that, using a cutoff value of 25 ng/ml, sPLA_2_-IIA is highly sensitive and specific in detecting sepsis^46^. Given that all deceased COVID-19 patients in our study had elevated sPLA_2_- IIA levels (>10 ng/ml), we propose incorporating sPLA_2_-IIA levels and specifically the newly identified PLA-BUN index into patient selection criteria to assess the efficacy of sPLA_2_-IIA inhibitors in increasing survival of severe COVID-19 patients.

In conclusion, we show for what we believe is the first time that sPLA_2_-IIA is a novel clinical indicator of COVID-19 disease severity and mortality and likely a central mechanistic driver of immune and multiorgan failure. Given the new stratification tools discovered here, sPLA_2_-IIA inhibitors merit testing in clinical trials against severe COVID-19 to reduce mortality.

## Methods

### Study Design

This retrospective study analyzed 127 plasma samples from patients hospitalized at Stony Brook University Medical Center (Stony Brook, NY, United States) from January to July 2020. This study followed Good Clinical Practice guidelines and was approved by the central institutional review board at Stony Brook University (IRB2020-00423). COVID-19 was diagnosed using the viral nucleic acid test (RT-PCR) per guidelines from Centers for Disease Control and Prevention (CDC). COVID-19 patients were classified into 3 groups: 1) mild = mild symptoms without pneumonia on imaging and discharged from inpatient care, 2) severe = respiratory tract or non-specific symptoms, pneumonia confirmed by chest imaging, oxygenation index below 94% on room air, and discharged from inpatient care, 3) deceased = expired during inpatient care.

### Sample Processing and Lipidomic Analyses

Frozen EDTA plasma samples were processed utilizing Biosafety Level 2 conditions as per CDC Guidelines for the handling and processing of specimens associated with Corona Virus Disease 2019. Metabolites were isolated from plasma via methanol-based containing 10 µl Splash Lipidomix (#330707, Avanti Polar Lipids, Alabaster, AL) and separated utilizing a reverse phase chromatography as previously described by Najdekr et al.^47^ Samples were analyzed utilizing an UHPLC-ESI-MS/MS system (UHPLC, Thermo Horizon Vanquish Duo System, MS, Thermo Exploris 480) and separation was achieved utilizing an Hypersil GOLD aQ UPLC column (100 x mm, 1.9 μm, Thermo Fisher Scientific, Part No. 25302-102130) with mobile phases composed of water containing 0.1% formic acid and methanol containing 0.1% formic acid. Metabolites were eluted over a 15 min gradient with the Exploris 480 operating in positive ion mode, utilizing an ion transfer tube temperature of 350 °C, sheath gas of 45, aux gas of 5, and spray voltage of 4000. MS data for all samples were collected using dynamic exclusion and then aligned with pooled samples collected using Thermo AquireX to achieve optimal metabolite identification in Lipid Search 4.0 and Thermo Compound Discoverer 2.3 software.

Targeted lipidomic analysis was performed using an Agilent 1200 HPLC tandem Thermo Quantum Ultra triple quadrupole mass spectrometer (Thermo Fisher Scientific, San Jose, USA) to quantify levels of major molecular species of lyso-phospholipids (lyso-PLs). C16, C18:1, C18:2, and C20:4 molecular species for lyso-phosphatidylcholine (lyso-PC), lyso- phosphatidylethanolamine (lyso-PE), and lyso-phosphatidylserine (lyso-PS) (Cayman Chemical, Ann Arbor, MI) were used as standards and deuterated Splash Lipidomix as internal standards. Lyso-PLs were separated using an Agilent Poroshell 120 EC-C18 1.9 µm (2.1×50 mm) with mobile phases composed of water containing 2 mM ammonium formate/0.1% formic acid (A) and methanol containing 1 mM ammonium formate/0.1% formic acid. Chromatographic gradient elution began at 40% A and remained there for the first minute, proceeding to 1% A at 6 minutes, staying there for 10.5 min before returning to 40% MPA over 1.5 min and remaining till the end of the 20 min run.

### sPLA_2_-IIA Concentrations

sPLA_2_-IIA levels in plasma were determined by ELISA (Cayman Chemical Company). Plasma samples were diluted (1:20-1:800) and assayed in duplicate. Concentrations of sPLA_2_-IIA in plasma were calculated using standard curves.

### Enzymatic Assay for sPLA_2_-IIA Activity

A subset of 34 patient samples (9 non-COVID-19, 8 mild, 7 severe, and 10 deceased COVID-19) were selected for PLA_2_ activity analysis. sPLA2 activity was assayed by modifying techniques from Kramer and Pepinsky^48^. Hydrolytic activity was determined in plasma samples from 34 patients (9 non-COVID-19, 8 mild, 7 severe, and 10 deceased COVID-19 patients) representing a wide range of sPLA_2_-IIA levels. Assays contained 5 µl of plasma in a final volume of 400 µl containing 50 mM Tris/NaCl, pH 8.5, with 5 mM CaCl_2_ and 5 nmol of 3H-oleate-labeled E. coli phospholipids and incubated for 30 mins at 37 °C.^2^ Lipids were extracted utilizing a modified Bligh and Dyer^49^, and hydrolyzed fatty acids were separated from phospholipids using thin layer chromatography (Silica Gel G) and a mobile phase of hexane:ether:formic acid (90:60:6, v:v:v), and visualized by iodine vapor relative to cold standards.

### Mitochonrial DNA Quantification

Mitochondrial DNA (mtDNA) was quantified in the same 34 patient samples as the enzymatic assay. Mitochondrial DNA (mtDNA) was quantified by adapting methods from Scozzi et. al.^31^ Using genes for human cytochrome C (MT-CYB) and cytochrome C oxidase subunit III (MT- COX3), mtDNA was quantified in plasma samples from the same 34 patients (9 non-COVID-19, 8 mild, 7 severe, and 10 deceased COVID-19 patients) as in the sPLA2 activity assay utilizing an ABI 7900HT real-time PCR instrument in 384-well format. Synthetic oligonucleotide copies of the MT-CYB and MT-COX3 genomic sequences (gBlock Gene Fragments from Integrated DNA Technologies) were included to generate a standard curve at 10^5^, 10^4^, 10^3^, and 10^2^ copies per µL. Primer sequences were as follows:

> forward MT-CYB: 5’– ATGACCCCAATACGCAAAA-3’
>
> reverse MT-CYB: 5’–CGAAGTTTCATCATGCGGAG-3’
>
> forward MT-COX3: 5’–ATGACCCACCAATCACATGC-3’
>
> reverse MT-COX3: 5’–ATCACATGGCTAGGCCGGAG-3’.

Each diluted serum sample was compared to a control reaction of a gBlock standard, and the delta-Ct was used to correct the calculated concentrations from triplicate reactions.

### Statistical Analyses

Untargeted lipidomic data were transformed, normalized, and analyzed using MetaboAnalyst 4.0. The Benjamini–Hochberg procedure was used to control the false discovery rate (FDR), and the molecules with FDR ≤ 0.1 and absolute log_2_ fold change (FC) ≥ 1.5 were considered as significant and biologically relevant. Individual metabolites, sPLA_2_ levels, sPLA_2_ activity, and mtDNA levels were compared between groups with non-parametric Mann-Whitney Wilcoxon tests at an α-level of 0.05. Spearman correlations between sPLA_2_ levels and clinical indices were computed in R. Receiver operating characteristic (ROC) curves, area under the curves (AUC), and confidence intervals were generated using the R packages ROCR and pROC.

### Decision Tree and Random Forest Construction

Eighty initial clinical indices were used as input variables to build a predictive model (i.e., decision tree) by recursive partitioning, using the Classification and Regression Trees (CART) algorithm^50^ implemented in the R package RPART. The tree model identified a set of predictive features (branch conditions) that best classified the 127 patients into the 4 groups: non-COVID-19, mild, severe, and deceased COVID-19 patients. The tree split points were determined by the Gini index with the minimum leaf size = 10. A tenfold cross-validation method was used to tune the tree model and evaluate its prediction accuracy. To avoid overfitting, the tree was pruned back to the smallest size while minimizing the cross-validated error. The classification accuracy of the tree to determine each group membership (e.g., deceased vs. non-deceased) was assessed using the area under the ROC curve. To further evaluate the relative feature importance in accurately separating severe and deceased COVID-19 patients, a random forest analysis was performed using the R package randomForest^51.^ An assembly of 1,000 random decision trees was constructed in each forest, and 10 forests were constructed in replicate. The importance of a given feature (i.e., one of the 80 clinical indices) was assessed by the decrease of prediction accuracy when such a feature was omitted in the model, based on two measurement metrics: Gini importance or Mean Decrease Impurity (MDI), and permutation importance or Mean Decrease Accuracy (MDA).

## Data Availability

The data that support the findings of this study are available from the corresponding author, FHC, upon request after publication of the manuscript.

## Acknowledgements

The authors wish to thank the Stony Brook Medicine Biobank for procuring and facilitating the distribution of plasma samples from COVID-19 and non-COVID hospitalized patients. We acknowledge the University of Arizona Data Science Institute for their help in applying machine learning and Dr. Manja Zec and Kirsten Lake for their constructive input to the manuscript.

This work was supported by the following grants from the National Institutes of Health: R01 AT008621 (FHC), R35 GM126922 (CEM), AI125770 (MD), AI136934 (MD), and P01-CA97132

(YAH). This work was also supported by the Veterans Affairs Merit Review Grant I01BX002924 (MD) and the Technology and Research Initiative Fund (BIO5 Institute, University of Arizona, FHC).

## Author Contributions

FHC conceived and designed the study. JY, RRK, MD, JMS., AJS, YAH, SS prepared IRB documentation and coordinated the collection of samples and clinical data. They also handled logistics of samples transfer between institutions. JMS, AJS, QW performed both untargeted and targeted metabolomics profiling and data analysis. MCS, SS performed sPLA_2_ activity assay. AJS performed sPLA_2_ ELISA. RS performed mitochondrial DNA analysis. XW, HZ, GY, BH, LJ, QW did the statistical analysis and modeling. FHC and JMS wrote the initial draft of the manuscript. CEM, XW, MD, YAH, AJS, CL contributed to text revisions and discussion.

## Conflict of Interest Statement

The authors declare no competing interest.

## Data Sharing

Anyone who wishes to share, reuse, remix, or adapt this material must obtain permission from the corresponding author.

## Notes

### Competing Interest Statement

The authors have declared no competing interest.

### Author Declarations

This study was approved by the central institutional review board at Stony Brook University (IRB2020-00423).

## References

1 Martins, R. et al. Disease Tolerance as an Inherent Component of Immunity. Annu Rev Immunol 37, 405–437, doi:10.1146/annurev-immunol-042718-041739 (2019).

2 Liao, D. et al. Haematological characteristics and risk factors in the classification and prognosis evaluation of COVID-19: a retrospective cohort study. Lancet Haematol 7, e671–e678, doi:10.1016/s2352-3026(20)30217-9 (2020).

3 Fajgenbaum, D. C. & June, C. H. Cytokine Storm. N Engl J Med 383, 2255–2273, doi:10.1056/NEJMra2026131 (2020).

4 Leisman, D. E. et al. Cytokine elevation in severe and critical COVID-19: a rapid systematic review, meta-analysis, and comparison with other inflammatory syndromes. Lancet Respir Med 8, 1233–1244, doi:10.1016/s2213-2600(20)30404-5 (2020).

5 Del Valle, D. M. et al. An inflammatory cytokine signature predicts COVID-19 severity and survival. Nat Med 26, 1636–1643, doi:10.1038/s41591-020-1051-9 (2020).

6 Sinha, P., Matthay, M. A. & Calfee, C. S. Is a “Cytokine Storm” Relevant to COVID-19? JAMA Intern Med 180, 1152–1154, doi:10.1001/jamainternmed.2020.3313 (2020).

7 Horby, P. et al. Dexamethasone in Hospitalized Patients with Covid-19 - Preliminary Report. N Engl J Med, doi:10.1056/NEJMoa2021436 (2020).

8 Mudd, P. A. et al. Distinct inflammatory profiles distinguish COVID-19 from influenza with limited contributions from cytokine storm. Sci Adv 6, doi:10.1126/sciadv.abe3024 (2020).

9 Remy, K. E. et al. Severe immunosuppression and not a cytokine storm characterizes COVID-19 infections. JCI Insight 5, doi:10.1172/jci.insight.140329 (2020).

10 Zheng, H. Y. et al. Elevated exhaustion levels and reduced functional diversity of T cells in peripheral blood may predict severe progression in COVID-19 patients. Cell Mol Immunol 17, 541–543, doi:10.1038/s41423-020-0401-3 (2020).

11 Pan, H. et al. Repurposed Antiviral Drugs for Covid-19 - Interim WHO Solidarity Trial Results. N Engl J Med, doi:10.1056/NEJMoa2023184 (2020).

12 Nienhold, R. et al. Two distinct immunopathological profiles in autopsy lungs of COVID-19. Nat Commun 11, 5086, doi:10.1038/s41467-020-18854-2 (2020).

13 Diao, B. et al. Reduction and Functional Exhaustion of T Cells in Patients With Coronavirus Disease 2019 (COVID-19). Front Immunol 11, 827, doi:10.3389/fimmu.2020.00827 (2020).

14 Shen, B. et al. Proteomic and Metabolomic Characterization of COVID-19 Patient Sera. Cell 182, 59-72.e15, doi:10.1016/j.cell.2020.05.032 (2020).

15 Wu, D. et al. Plasma metabolomic and lipidomic alterations associated with COVID-19. National Science Review 7, 1157–1168, doi:10.1093/nsr/nwaa086 (2020).

16 Dore, E. & Boilard, E. Roles of secreted phospholipase A(2) group IIA in inflammation and host defense. Biochim Biophys Acta Mol Cell Biol Lipids 1864, 789–802, doi:10.1016/j.bbalip.2018.08.017 (2019).

17 van Hensbergen, V. P., Wu, Y., van Sorge, N. M. & Touqui, L. Type IIA Secreted Phospholipase A2 in Host Defense against Bacterial Infections. Trends Immunol 41, 313–326, doi:10.1016/j.it.2020.02.003 (2020).

18 Chung, K. P. et al. Increased Plasma Acetylcarnitine in Sepsis Is Associated With Multiple Organ Dysfunction and Mortality: A Multicenter Cohort Study. Crit Care Med 47, 210–218, doi:10.1097/ccm.0000000000003517 (2019).

19 Ruiz, M. et al. Circulating acylcarnitine profile in human heart failure: a surrogate of fatty acid metabolic dysregulation in mitochondria and beyond. Am J Physiol Heart Circ Physiol 313, H768–h781, doi:10.1152/ajpheart.00820.2016 (2017).

20 Ceriello, A. Hyperglycemia and the worse prognosis of COVID-19. Why a fast blood glucose control should be mandatory. Diabetes Res Clin Pract 163, 108186, doi:10.1016/j.diabres.2020.108186 (2020).

21 Hariyanto, T. I. & Kurniawan, A. Anemia is associated with severe coronavirus disease 2019 (COVID-19) infection. Transfus Apher Sci 59, 102926, doi:10.1016/j.transci.2020.102926 (2020).

22 Schwarz, B. et al. Cutting Edge: Severe SARS-CoV-2 Infection in Humans Is Defined by a Shift in the Serum Lipidome, Resulting in Dysregulation of Eicosanoid Immune Mediators. J Immunol 206, 329–334, doi:10.4049/jimmunol.2001025 (2021).

23 Barberis, E. et al. Large-Scale Plasma Analysis Revealed New Mechanisms and Molecules Associated with the Host Response to SARS-CoV-2. Int J Mol Sci 21, doi:10.3390/ijms21228623 (2020).

24 Vadas, P. Elevated plasma phospholipase A2 levels: correlation with the hemodynamic and pulmonary changes in gram-negative septic shock. J Lab Clin Med 104, 873–881 (1984).

25 Guidet, B. et al. Secretory non-pancreatic phopholipase A2 in severe sepsis: relation to endotoxin, cytokines and thromboxane B2. Infection 24, 103–108, doi:10.1007/bf01713312 (1996).

26 Vadas, P., Pruzanski, W. & Farewell, V. A predictive model for the clearance of soluble phospholipase A2 during septic shock. J Lab Clin Med 118, 471–475 (1991).

27 Waydhas, C. et al. Inflammatory Mediators, Infection, Sepsis, and Multiple Organ Failure After Severe Trauma. Archives of Surgery 127, 460–467, doi:10.1001/archsurg.1992.01420040106019 (1992).

28 Kitadokoro, K., Hagishita, S., Sato, T., Ohtani, M. & Miki, K. Crystal structure of human secretory phospholipase A2-IIA complex with the potent indolizine inhibitor 120-1032. J Biochem 123, 619–623, doi:10.1093/oxfordjournals.jbchem.a021982 (1998).

29 Bjørndal, B. et al. Associations between fatty acid oxidation, hepatic mitochondrial function, and plasma acylcarnitine levels in mice. Nutrition & Metabolism 15, 10, doi:10.1186/s12986-018-0241-7 (2018).

30 McGill, M. R. et al. Circulating acylcarnitines as biomarkers of mitochondrial dysfunction after acetaminophen overdose in mice and humans. Archives of Toxicology 88, 391–401, doi:10.1007/s00204-013-1118-1 (2014).

31 Scozzi, D. et al. Circulating mitochondrial DNA is an early indicator of severe illness and mortality from COVID-19. JCI Insight, doi:10.1172/jci.insight.143299 (2021).

32 Ayres, J. S. A metabolic handbook for the COVID-19 pandemic. Nature Metabolism 2, 572–585, doi:10.1038/s42255-020-0237-2 (2020).

33 Maitra, U., Chang, S., Singh, N. & Li, L. Molecular mechanism underlying the suppression of lipid oxidation during endotoxemia. Mol Immunol 47, 420–425, doi:10.1016/j.molimm.2009.08.023 (2009).

34 Cheng, A. et al. Diagnostic performance of initial blood urea nitrogen combined with D-dimer levels for predicting in-hospital mortality in COVID-19 patients. Int J Antimicrob Agents 56, 106110, doi:10.1016/j.ijantimicag.2020.106110 (2020).

35 Boudreau, L. H. et al. Platelets release mitochondria serving as substrate for bactericidal group IIA-secreted phospholipase A2 to promote inflammation. Blood 124, 2173–2183, doi:10.1182/blood-2014-05-573543 (2014).

36 Hurt-Camejo, E., Camejo, G., Peilot, H., Oörni, K. & Kovanen, P. Phospholipase A(2) in vascular disease. Circ Res 89, 298–304, doi:10.1161/hh1601.095598 (2001).

37 Murakami, M. et al. The Roles of the Secreted Phospholipase A(2) Gene Family in Immunology. Adv Immunol 132, 91–134, doi:10.1016/bs.ai.2016.05.001 (2016).

38 Chilton, F. Would the real role(s) for secretory PLA2s please stand up. J Clin Invest 97, 2161–2162, doi:10.1172/jci118654 (1996).

39 Atsumi, G. et al. The perturbed membrane of cells undergoing apoptosis is susceptible to type II secretory phospholipase A2 to liberate arachidonic acid. Biochim Biophys Acta 1349, 43–54, doi:10.1016/s0005-2760(97)00082-9 (1997).

40 Roh, J. S. & Sohn, D. H. Damage-Associated Molecular Patterns in Inflammatory Diseases. Immune Netw 18, e27, doi:10.4110/in.2018.18.e27 (2018).

41 Tan, T. L. & Goh, Y. Y. The role of group IIA secretory phospholipase A2 (sPLA2-IIA) as a biomarker for the diagnosis of sepsis and bacterial infection in adults-A systematic review. PLoS One 12, e0180554, doi:10.1371/journal.pone.0180554 (2017).

42 Anderson, B. O., Moore, E. E. & Banerjee, A. Phospholipase A2 regulates critical inflammatory mediators of multiple organ failure. J Surg Res 56, 199–205, doi:10.1006/jsre.1994.1032 (1994).

43 Corke, C., Glenister, K. & Watson, T. Circulating secretory phospholipase A2 in critical illness--the importance of the intestine. Crit Care Resusc 3, 244–249 (2001).

44 Zeiher, B. G. et al. LY315920NA/S-5920, a selective inhibitor of group IIA secretory phospholipase A2, fails to improve clinical outcome for patients with severe sepsis. Crit Care Med 33, 1741–1748, doi:10.1097/01.ccm.0000171540.54520.69 (2005).

45 Abraham, E. et al. Efficacy and safety of LY315920Na/S-5920, a selective inhibitor of 14-kDa group IIA secretory phospholipase A2, in patients with suspected sepsis and organ failure. Crit Care Med 31, 718–728, doi:10.1097/01.Ccm.0000053648.42884.89 (2003).

46 Berg, E. et al. Measurement of a Novel Biomarker, Secretory Phospholipase A2 Group IIA as a Marker of Sepsis: A Pilot Study. Journal of Emergencies, Trauma, and Shock 11, 135, doi:10.4103/JETS.JETS_29_17 (2018).

47 Najdekr, L., Blanco, G. R. & Dunn, W. B. Collection of Untargeted Metabolomic Data for Mammalian Urine Applying HILIC and Reversed Phase Ultra Performance Liquid Chromatography Methods Coupled to a Q Exactive Mass Spectrometer. Methods Mol Biol 1996, 1–15, doi:10.1007/978-1-4939-9488-5_1 (2019).

48 Kramer, R. M. & Pepinsky, R. B. Assay and purification of phospholipase A2 from human synovial fluid in rheumatoid arthritis. Methods Enzymol 197, 373–381, doi:10.1016/0076-6879(91)97163-s (1991).

49 Bligh, E. G. & Dyer, W. J. A rapid method of total lipid extraction and purification. Can J Biochem Physiol 37, 911–917, doi:10.1139/o59-099 (1959).

50 Breiman, L., Friedman, J., Olshen, R. & Stone, C. Classification and Regression Trees. (Chapman & Hall, 1984).

51 Breiman, L. Random Forests. Machine Learning 45, 5–32, doi:10.1023/A:1010933404324 (2001).

