## Supplemental Figures 1-3 for "Group IIA Secreted Phospholipase A_2_ Plays a Central Role in the Pathobiology of COVID-19"

### SUPPLEMENTARY FIGURES

#### Figure S1

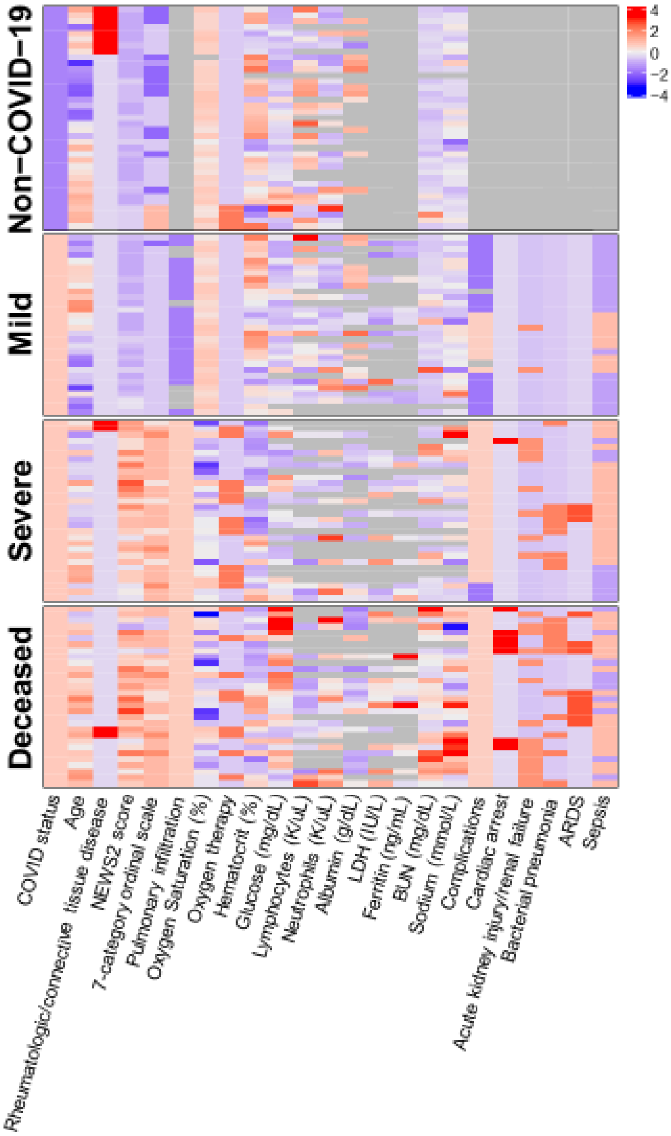


**Figure S1. Clinical Indices and Disease Status.** Clinical indices that vary significantly (FDR<0.05, F-test) across 4 patient groups are shown in a heatmap, with blue to red representing low to high values of each index, and the color intensity representing the magnitude of value (mean-centered, scaled by the standard deviation, and log-transformed with non-Gaussian distribution). Missing values are shown in grey.

### Figure S2

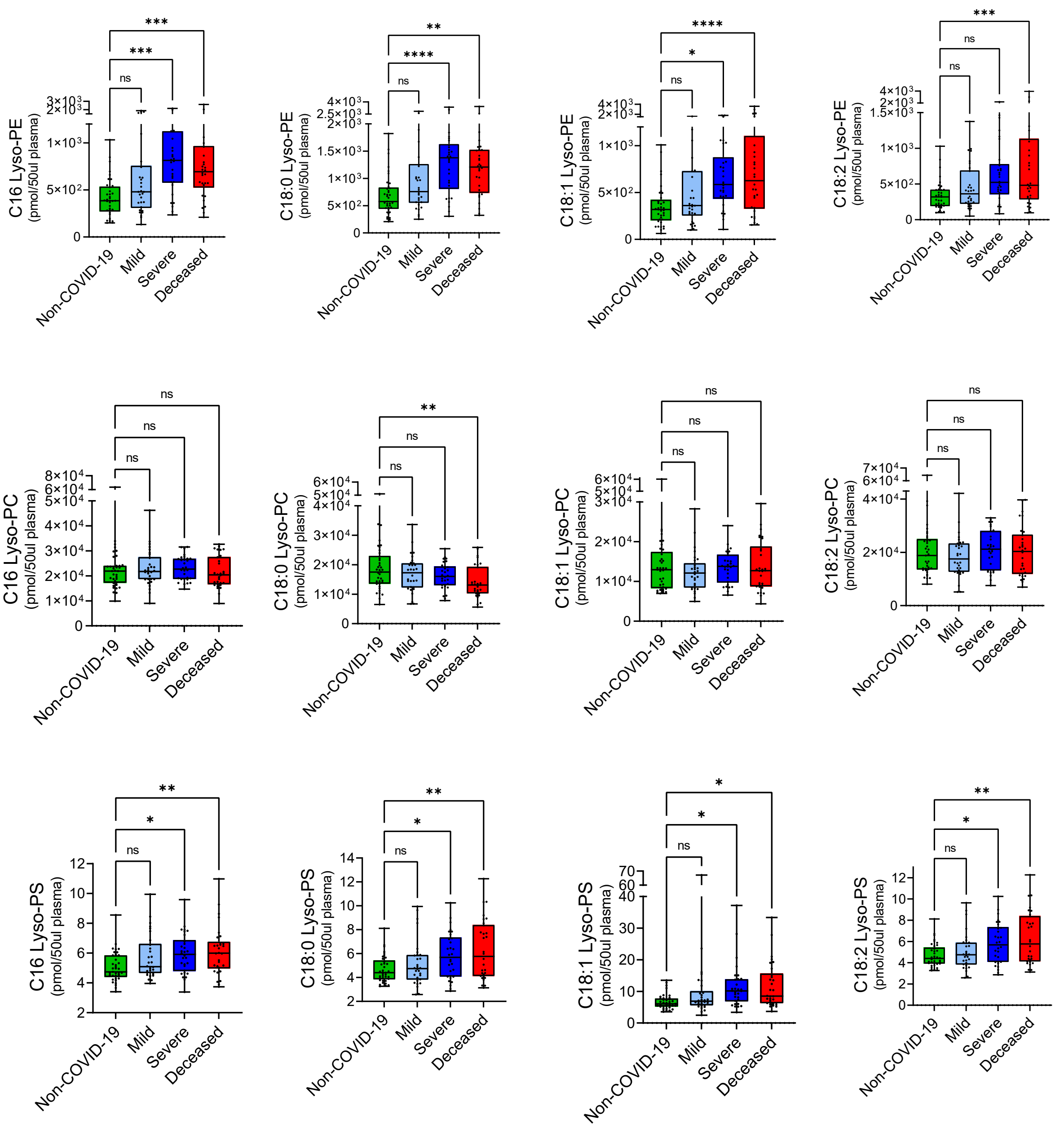


**Figure S2. Targeted Lyso-phospholipid (Lyso-PL) Analysis.** Lyso-PLs were identified as top molecules of interest from the qualitative untargeted lipidome data set. Samples were re-analyzed utilizing lyso-PL standards in a targeted lyso-PL method. Asterisks indicate significance by ANOVA (1-way ANOVA with Dunnen multiple comparison): * $p<0.05$; ** $p<0.01$; *** $p<0.001$; **** $p<0.0001$

**Figure S3**

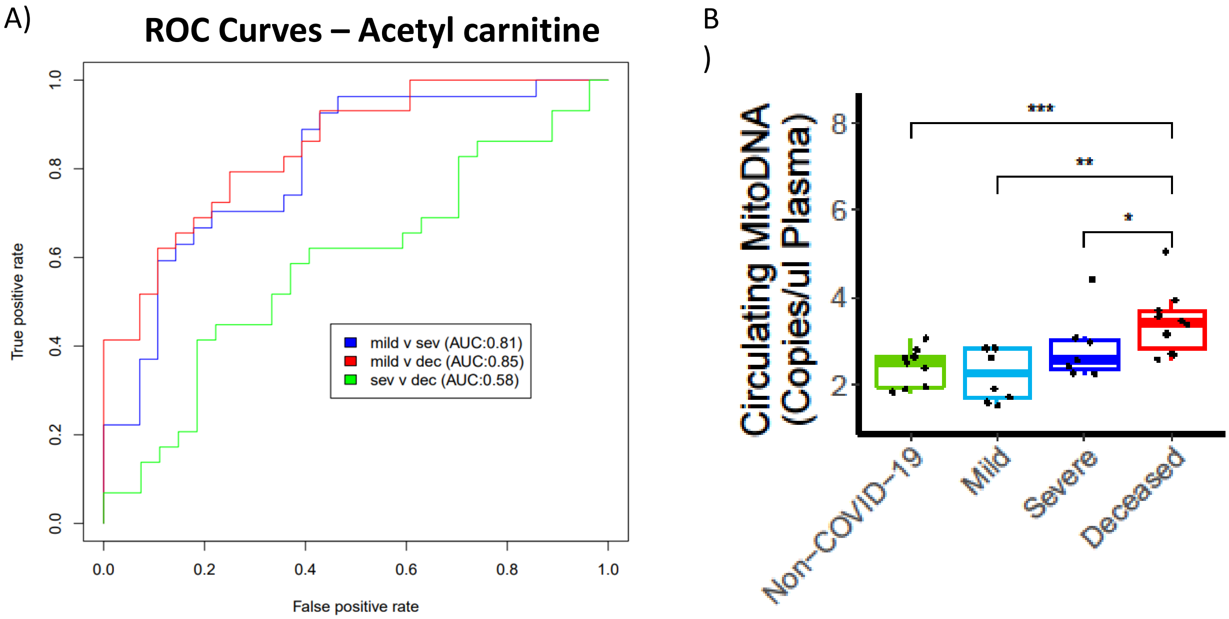


**Figure S3. Markers of Mitochondrial Dysfunction.** A) ROC curves demonstrate the performance of acetylcarnitine as a predictor of disease status. B) Mitochondrial DNA was quantified in a selected subset of patients (n=34; 9 non-COVID-19, 8 mild, 7 severe, and 10 deceased COVID-19 patients). Summed copy numbers of genes for human cytochrome C and cytochrome C oxidase subunit III are shown. Data are log transformed with one-sided Wilcoxon test significance indicated as: * p<0.05; ** p<0.01; *** p<0.001.
